# Health Data Reporting Challenges in the Age of DHIS2: a Mixed-Methods Review in the Teso Sub-Region of Uganda

**DOI:** 10.1101/2024.09.18.24313921

**Authors:** Margaret R. Lawrence, Benjamin P. Fuller, Herbert Kirya Isabirye, Carol Kyozira, Issa Makumbi, Andrew Bakainaga, Christopher C. Moore, Richard Ssekitoleko

## Abstract

In Uganda, the complete and timely reporting of priority disease data by health centers is critical for detecting outbreaks. However, health centers face significant reporting challenges, which compromise data fidelity and public health. To assess the efficacy of existing systems, we aimed to (1) compare reporting performance among districts in the Teso region of Eastern Uganda, (2) ascertain the association between volume of outpatient department (OPD) visits and reporting rates at facilities, and (3) characterize reporting challenges. We queried Uganda’s Electronic Health Management Information System database, DHIS2, regarding the completeness (reporting rate) and timeliness (reporting rate on time) of priority disease data submission within Teso from January-April 2024. We selected the 3 lowest-performing districts in both metrics and assessed reporting at all health centers within these districts. We used linear regression to determine the relationship between the aggregate number of outpatient visits per health facility and their mean reporting rates. At 7 health facilities across the 3 districts, we interviewed staff to enumerate reporting challenges. Of all 11 districts in Teso, 7 (64%) scored >80% for completeness and 2 (18%) scored >80% for submission timeliness across health facilities. The median (IQR) scores for completeness and timeliness were 83% (75-93%) and 69% (55-75%), respectively. The 3 lowest performing districts in both metrics were Soroti City, Soroti district, and Ngora district. In Soroti City, increasing number of OPD visits was associated with higher reporting rate percentages (b [slope]=0.005, p=0.01). The main reasons given for low reporting rates included limited staff trained on data entry protocols, competing responsibilities among reporting personnel, and lack of reliable internet access. The implementation of DHIS2 has improved public health and disease surveillance in Uganda, but institutional-level data reporting challenges continue to impair disease tracking. Targeted interventions to relieve such barriers is possible.

## INTRODUCTION

The complete and timely reporting of health data by health system constituents is critical for facilitating disease surveillance, outbreak detection, and response activities.^1^ Data that are incomplete, delayed, or have not been validated do not effectively capture real-time disease trends and may lead to biased, uninformed decision-making at both the point of generation of the data and at the national level.^1,2^ It is thus essential to examine data reporting processes and challenges of such so that data quality and fidelity may be optimized and resources properly allocated.

A cornerstone of Uganda’s public health framework, the World Health Organization’s Integrated Disease Surveillance and Response (IDSR) strategy was first adapted by Uganda in 2001 with the aims of monitoring disease trends and promoting rapid interventions to minimize the impact of public health threats across the country.^3^ As part of IDSR implementation, all district-level health centers are required to report pertinent health statistics on an immediate, weekly, monthly, or quarterly basis depending on the urgency and implications of the data^4^. Uganda has identified a list of priority diseases of outbreak potential, known as 033b data, to be reported by all health facilities on a weekly basis.^4^ Diseases included on the 033b form include malaria, dysentery, severe acute respiratory illness, acute flaccid paralysis, adverse events following immunization, animal bites (suspected rabies), bacterial meningitis, cholera, Guinea worm disease (dracunculiasis), measles, neonatal tetanus, plague, typhoid fever, hepatitis B, rifampicin-resistant tuberculosis cases, yellow fever, viral hemorrhagic fever syndromes (Ebola, Marburg, Lassa fever, Crimean-Congo), leprosy, anthrax, maternal death, macerated stillbirths, fresh stillbirths, and early neonatal deaths (0-7 days)^4,5^. For each disease category, the reporting person is to indicate the number of new cases detected during the week, the number of deaths that occurred during the week attributable to the condition, those cases that were tested, and those that yielded positive results.^5^

Information from 033b reports ultimately reaches the Health-Sub-District Headquarters and District Health Officer through a hierarchical reporting system that begins with focal surveillance and medical records officers at Health Center levels II, III, IV, and hospitals.^4–6^ Once reported from the health facilities by means of a short message service (SMS) reporting system known as mobile tracking (mTrac), data are compiled by district biostatisticians and submitted to the national electronic health information system platform, District Health Information System 2 (DHIS2), as aggregate data.^4,6,7^ This platform assesses individual health centers within each district on the bases of timely (by midday every Monday following the week of the relevant reporting period) and complete (by midday every Wednesday following the week of the relevant reporting period) data reporting. To ensure accurate and informed surveillance at the national level, the WHO recommends that each district produces a minimum of 80% of the required reports, with 80% being submitted on time.^6,8^

Despite this national target, there continue to be challenges across the country with data completeness, timeliness, and accuracy, with all institutional levels of surveillance struggling to meet recommended targets.^1,6,9,10^ Suggested barriers have included little to no coordination between those collecting the data, those who analyze it, and those who use it for decision-making as well as a lack of prioritization of surveillance in light of budget and human-resource constraints.^11^ With technological advances in IDSR– including the implementation of mTrac and DHIS2– have also come primarily technological challenges, including limited access to wireless networks contributing to delay in data submission and errors in manual reporting processes.^6^

To assess the efficacy of existing surveillance systems, it is critical to identify the factors contributing to incomplete and delayed data reporting as well as those areas where data quality and accuracy may be compromised. In this qualitative mixed-methods review, we aimed to assess weekly 033b reporting performance among districts in the rural Teso sub-region of Eastern Uganda and to illuminate facilities’ barriers to complete and timely data reporting to better optimize national surveillance of and response to disease outbreaks. Previous studies have analyzed and compared timeliness and completeness of weekly surveillance data across reporting regions of Uganda, but to our knowledge this is the first study in the country that seeks to characterize the unique challenges of data reporting through standardized interviews with reporters themselves.^6,11,12^

## METHODS

### Study setting

Uganda has a decentralized public health system with health services overseen at the district, regional, and national levels.^13,14^ Working at the community level are Village Health Teams, or Community Health Workers, individuals who closely engage with community members and provide on-the-ground disease surveillance, health promotion, treatment and referral, and voluntary services.^13,15,16^ District health teams and the Ministry of Health manage health services across the country’s 146 districts.^13^ Almost all districts have a General Hospital and Health Centers (HCs) levels II-IV. Staffing and service capacity increase from HC level II-IV, with HC IVs serving populations of up to 100,000 people and overseeing the level II and III centers within the district.^13^ There are also regional and national referral hospitals throughout the country.^14,13^ Within the sub-region of Teso, there are ten reporting districts, including Serere, Kumi, Amuria, Kaberamaido, Bukedea, Kapelebyong, Kalaki, Katawki, Soroti, and Ngora districts.^17^ Soroti City is considered a separate reporting entity within Soroti district.^17^ As of 2018, there were 241 documented health facilities across the eleven reporting entities, each of which is required to submit weekly priority disease surveillance data.^17^

### Data abstraction and study design

We queried the nationwide Uganda Electronic Health Management Information System (eHMIS) database, DHIS2, regarding the completeness (reporting rate) and timeliness (reporting rate on time) of required weekly submission of 033b (the priority disease tracking form) within the Teso sub-region from January through April 2024. Per the WHO guidelines, we deemed those districts meeting ≥80% in both metrics to have met the requirements. From this preliminary analysis, we then selected the three lowest-performing districts and queried all health centers within these districts to assess for rates of completeness and timeliness by facility. For ethical reasons, we deidentified all health facilities within the three districts and labeled them with letters of the alphabet. This study was conducted as part of Uganda Ministry of Health quality improvement projects and thus did not require any approval from a research ethics committee.

To establish reasons for poor reporting within the three lowest-performing districts, we visited their district health offices to speak with district health team members at each. Study personnel, accompanied by either a health information officer or a district surveillance focal person from the respective district, then visited seven of the 54 total health centers across the three districts.

Those selected for interview ranged from poorest to well-performing in their districts; this was in an effort to better understand potential differences between facilities and varied approaches to managing shared challenges at different health centers from level II-level IV. We excluded private sector health facilities.

To guide discussions with health facility personnel, we developed a standardized questionnaire (Supplementary Figure 1) to assess the barriers to complete and timely reporting of priority diseases to the DHIS2 system and whether there were any difficulties submitting particular data elements to DHIS2. We analyzed the relationship between facilities’ volume of monthly outpatient department (OPD) visits and the completeness of their 033b reporting to better understand other potential factors associated with low reporting rates of 033b data.

### Statistical analysis

We evaluated rates of weekly 033b data submission at the district level using the DHIS2 data visualizer, a tool accessible through the Uganda eHMIS from January-April, 2024. We determined the median and interquartile ranges (IQRs) of the scores. We considered percentages ≥80% in each category as satisfactory for both measures of timeliness and completeness. Using the data outputs from DHIS2, we stratified district performance and identified the 3 lowest-performing districts. We then studied the completeness and timeliness of reporting by individual health facilities within these 3 districts and summarized the mean percentages, and their median and IQRs in tables. We used linear regression analysis to understand the effect of volume of OPD visits reported by individual health facilities on their average 033b reporting rates. OPD attendance was the independent (x) variable and reporting completeness was the dependent (y) variable. Facilities that did not report OPD attendance for one or more months were excluded from the regression analyses altogether. We considered p values <0.05 to be statistically significant.

### Qualitative analysis

We conducted focused interviews with district health teams across the three lowest-performing districts (Soroti district, Soroti City, and Ngora district). This was followed by physical visits to seven selected facilities (health centers C and I in Soroti district; facilities D, O, and T in Soroti City; and health centers C and H in Ngora district) where we conducted standardized key-informant interviews with 21 medical personnel, including HMIS focal persons, medical records assistants, health information assistants, district biostatisticians, clinicians, nurses, and a district health officer. In general, HCs were selected given their low performance in both complete and timely data reporting metrics as abstracted from DHIS2; however, 2 of the 7 participating health centers consistently reported above target, including health center D in Soroti City and health center C in Ngora. By visiting these higher performing facilities, we were able to better compare and contrast challenges faced by health centers and learn how shared obstacles are differentially approached by underreporting and successfully-reporting institutions.

## RESULTS

### Completeness and timeliness of 033b data reporting by district in the Teso sub-region, January-April, 2024

Of the 11 districts in Teso, 4 (36%) had a mean reporting rate <80%, including Kalaki (79%), Soroti City (71%), Soroti district (64%), and Ngora (63%). The remaining 7 (64%) districts met or exceeded the target for completeness: Bukedea (83%), Serere (100%), Amuria (95%), Kapelebyong (89%), Kaberamaido (91%), Katawki (81%), and Kumi districts (90%). Among the 11 districts, 2 (18%) met the ≥80% timeliness target of 033b data submission, including Serere (99%) and Kapelebyong (88%). The median (IQR) scores for completeness and timeliness were 83% (71-91%) and 69% (53-79%), respectively (Tables 1 and 2 and Figure 1). Soroti district, Soroti City, and Ngora district were the 3 lowest-performing districts in both complete and timely metrics, and were selected for further analysis.

**Figure 1.**
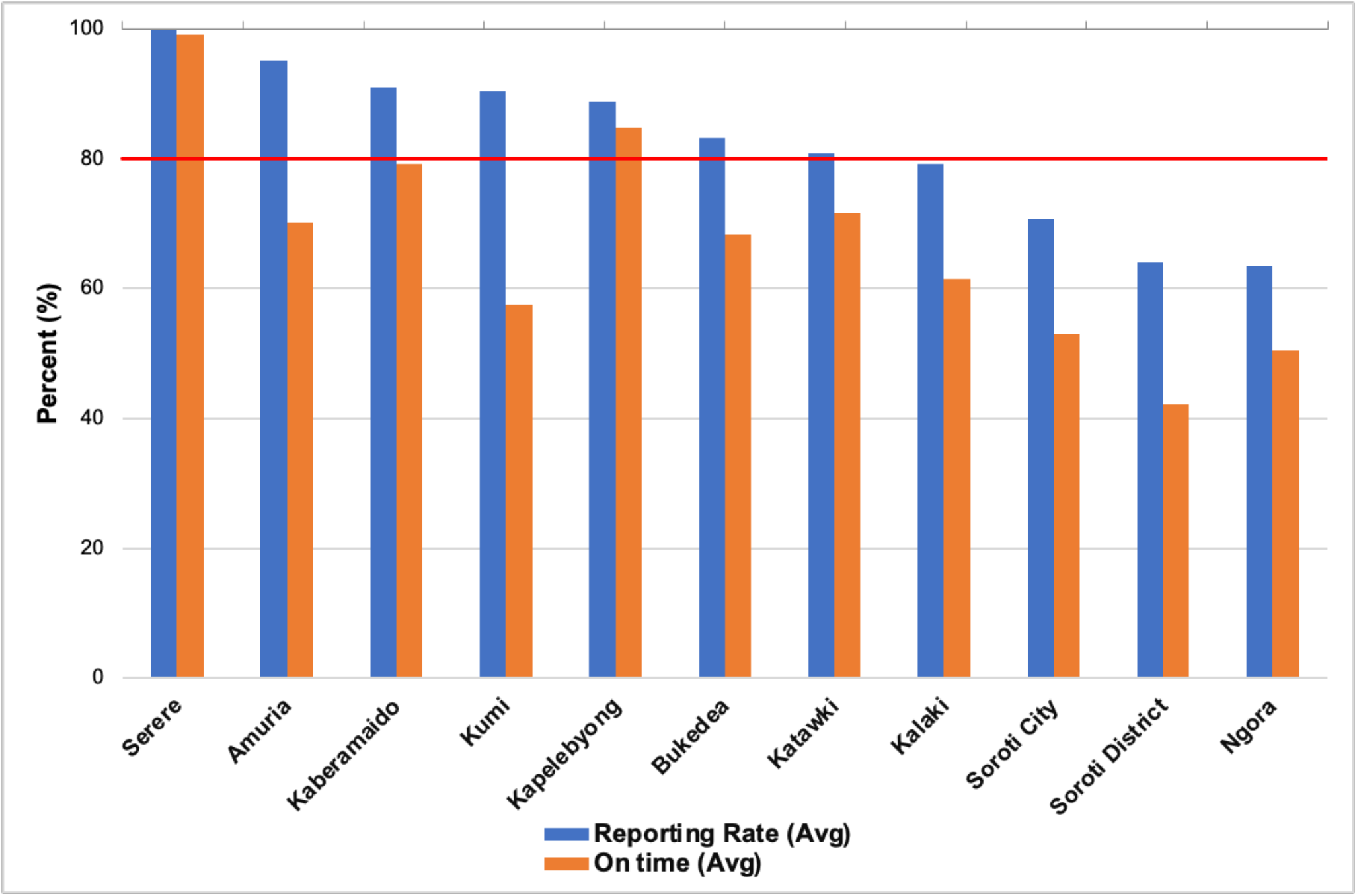
Average rates of complete and timely reporting of 033b data by district in the Teso region, January-April, 2024. The red line represents the cutoff for the WHO recommendation that each district produce at least 80% of required reports, with 80% submitted on time.

**Table 1.** Rates of complete reporting of 033b data by district in the Teso region of Uganda, January-April, 2024.

| Organizational Unit | Monthly scores for completeness (%) |  |  |  | Mean |
| --- | --- | --- | --- | --- | --- |
| Teso Districts | January | February | March | April |  |
| Serere | 100 | 100 | 100 | 100 | 100 |
| Amuria | 98 | 100 | 92 | 90 | 95 |
| Kaberamaido | 92 | 100 | 100 | 72 | 91 |
| Kumi | 81 | 84 | 99 | 97 | 90 |
| Kapelebyong | 95 | 100 | 91 | 70 | 89 |
| Bukedea | 100 | 100 | 63 | 70 | 83 |
| Katakwi | 91 | 100 | 64 | 69 | 81 |
| Kalaki | 94 | 100 | 54 | 69 | 79 |
| Soroti City | 72 | 79 | 65 | 67 | 71 |
| Soroti district | 83 | 72 | 52 | 50 | 64 |
| Ngora | 79 | 81 | 50 | 44 | 64 |
| **Median (IQR) aggregate score for Teso Region** | --- | -- | -- | -- | **83 (71-91)** |

**Table 2.** Rates of timely reporting of 033b data by district in the Teso region of Uganda, January-April, 2024.

| **Organizational Unit** | **Monthly scores for timeliness (%)** |  |  |  | **Mean** |
| --- | --- | --- | --- | --- | --- |
| Teso Districts | January | February | March | April |  |
| Serere | 100 | 100 | 97 | 100 | 99 |
| Amuria | 75 | 89 | 57 | 90 | 70 |
| Kaberamaido | 84 | 95 | 66 | 72 | 79 |
| Kumi | 48 | 47 | 65 | 70 | 58 |
| Kapelebyong | 95 | 100 | 77 | 68 | 85 |
| Bukedea | 73 | 98 | 50 | 54 | 69 |
| Katakwi | 82 | 99 | 49 | 57 | 72 |
| Kalaki | 73 | 88 | 35 | 50 | 62 |
| Soroti City | 55 | 59 | 47 | 51 | 53 |
| Soroti district | 53 | 53 | 34 | 28 | 42 |
| Ngora | 61 | 67 | 36 | 36 | 50 |
| **Median (IQR) aggregate score for Teso Region** | -- | -- | -- | -- | **69 (53-79)** |

### Completeness and timeliness of 033b data reporting by health facilities within Soroti district, Soroti City, and Ngora district, January-April, 2024

Of the 16 reporting facilities in Soroti district, 3 (19%) scored a mean ≥80% for the completeness of weekly 033b reporting, including facilities A (94%), B (88%), and C (81%). There was 1 (6%) of 16 reporting health facilities, facility A, that scored on average ≥80% for timeliness (88%) (Figure 2). The median (IQR) scores for completeness and timeliness were 66% (47-75%) and 47% (19-59.5%), respectively (Supplementary Tables 1 and 2). Among the 22 reporting health facilities in Soroti City, 13 (59%) scored on average ≥80% for completeness of weekly 033b reporting (Figure 3). There were 2 (9%) centers that reported on average ≥80% for timeliness, including facility G (81%) and facility M (81%) (Figure 3). Completeness and timeliness median (IQR) scores were 81% (56-88%) and 56% (34.5-75%), respectively (Supplementary Tables 3 and 4). Of the 13 reporting facilities in Ngora, 5 (38%) scored on average ≥80% for completeness of weekly 033b reporting, including facilities A, B, C, D, and E (81%). Only 1 (8%) of 13 health facilities, facility C (81%), scored on average ≥80% timeliness (Figure 4). The median (IQR) scores were 69% (53-81%) and 56% (34.5-69%) for completeness and timeliness, respectively (Supplementary Tables 5 and 6).

**Figure 2.**
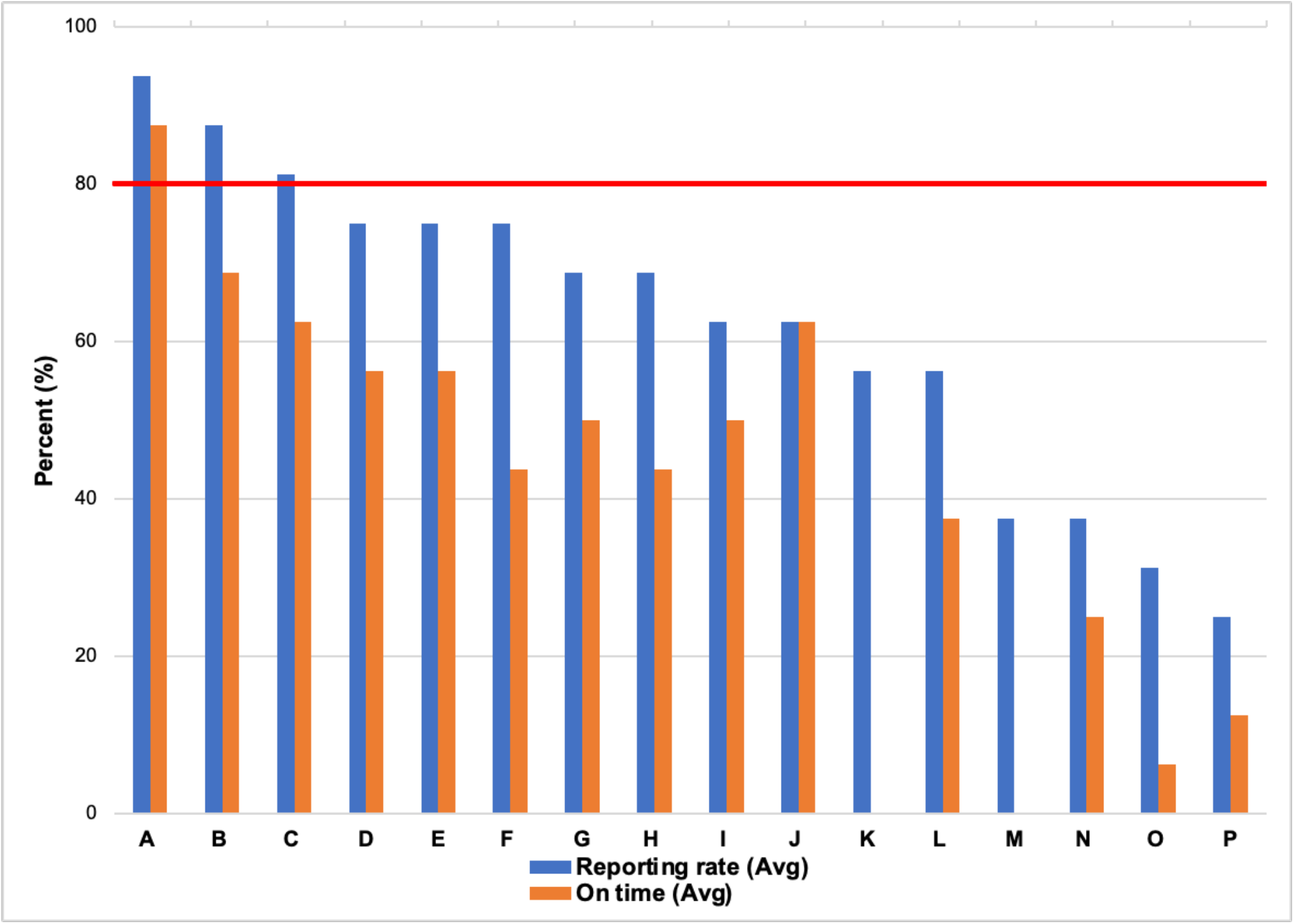
Average rates of complete and timely reporting of 033b data by health centers in Soroti district, first four months of 2024. The red line represents the cutoff for the WHO recommendation that each district produce at least 80% of required reports, with 80% submitted on time.

**Figure 3.**
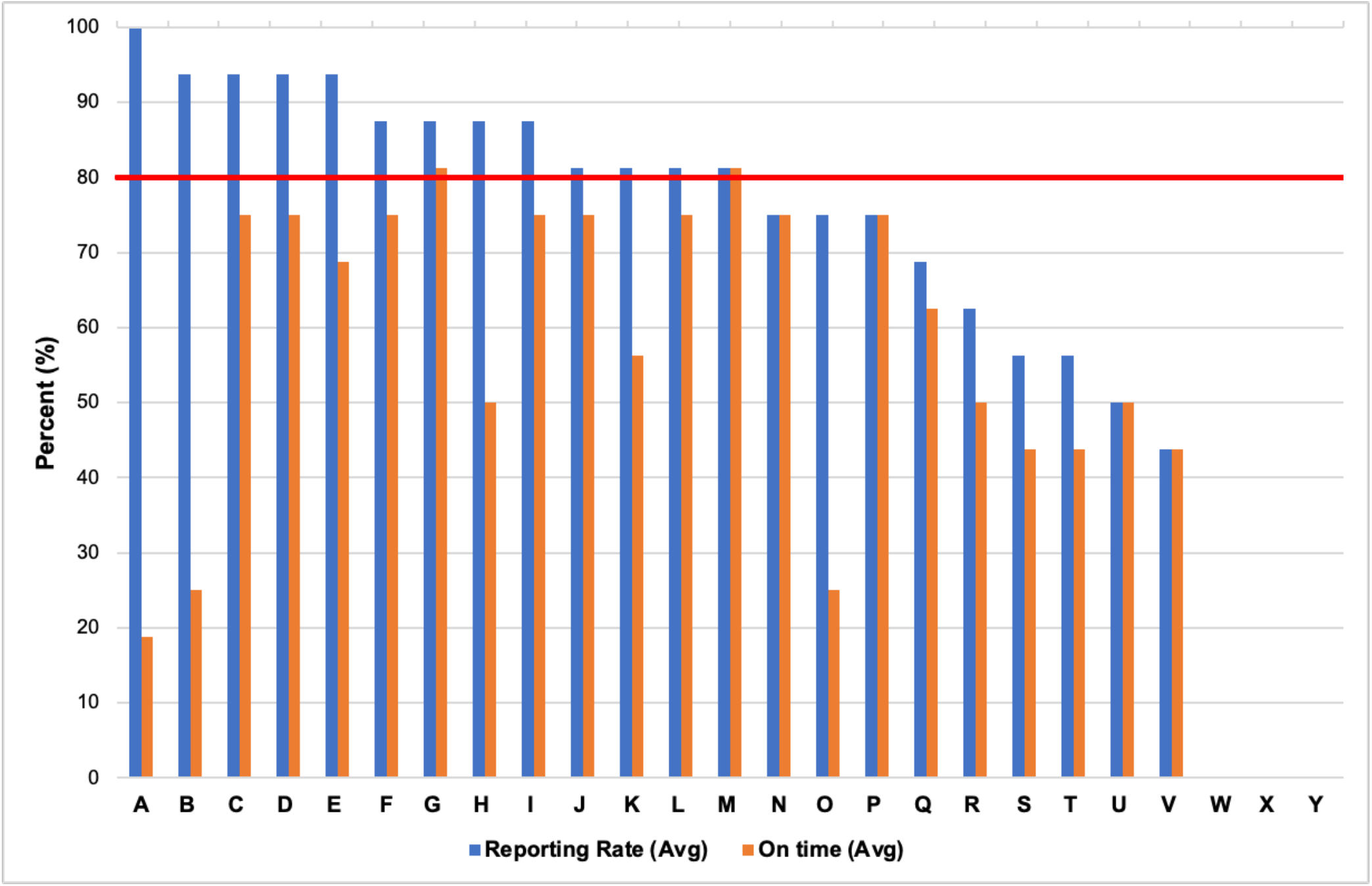
Average rates of complete and timely reporting of 033b data by health facilities within Soroti City, first four months of 2024. The red line represents the cutoff for the WHO recommendation that each district produce at least 80% of required reports, with 80% submitted on time.

**Figure 4.**
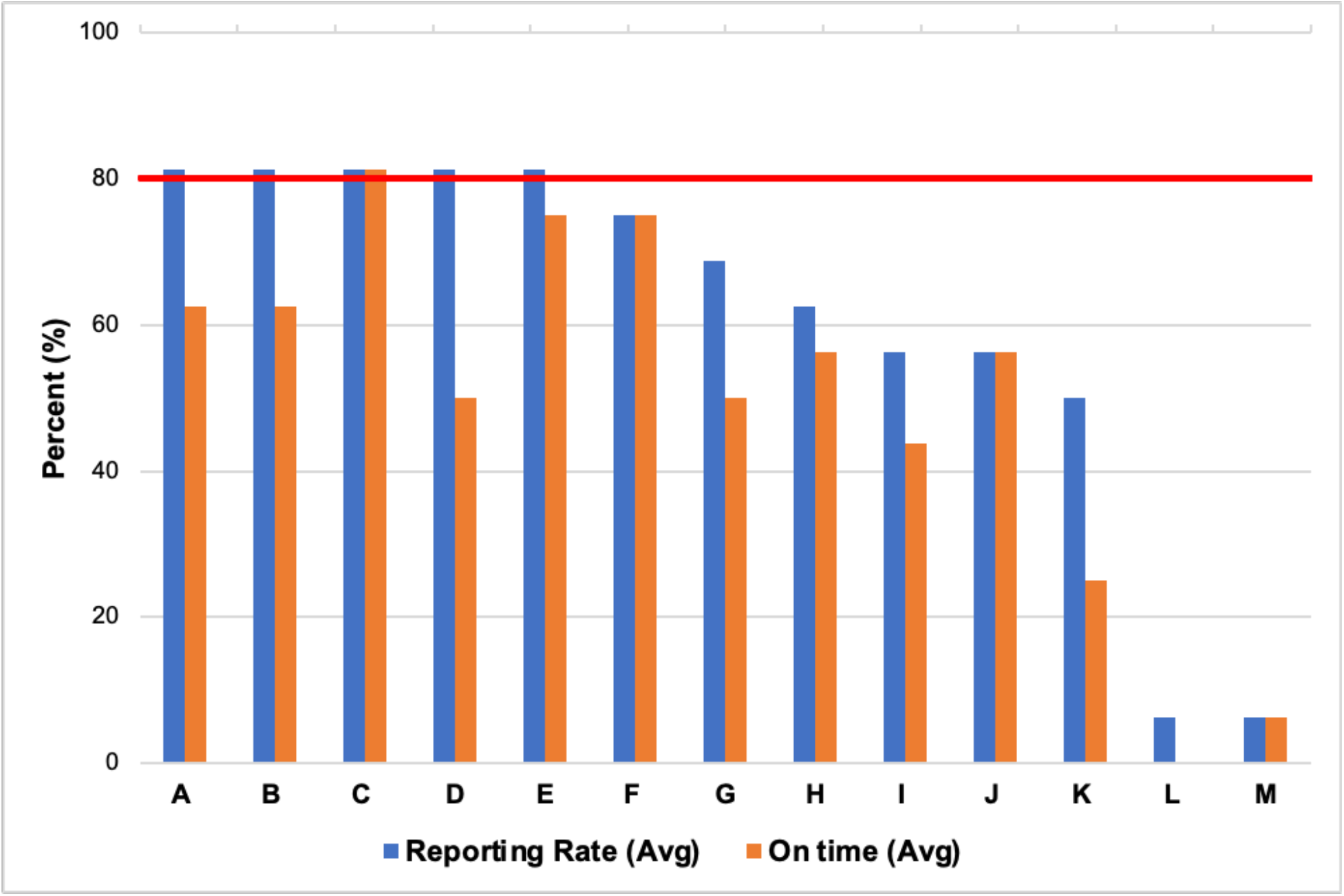
Average rates of complete and timely reporting of 033b data by health facilities within Ngora district, first four months of 2024. The red line represents the cutoff for the WHO recommendation that each district produce at least 80% of required reports, with 80% submitted on time.

### Linear Regression Analyses; aggregate OPD attendance and average 033b reporting rate for Soroti district, Soroti City, and Ngora district January-April, 2024

There was no association between volume of OPD visits (Supplementary Tables 7 and 8) and reporting rate percentages (completeness) for Soroti or Ngora districts. In Soroti City, there was a statistically significant (b [slope]=0.005, p=0.01) positive correlation between the number of OPD visits (Supplementary Table 9) and completeness of 033b data reporting over the study period.

### Major and minor themes from key informant interviews

Three of the 7 health centers, including health centers D, O, and T in Soroti City and health center H in Ngora, cited limited staffing, competing responsibilities among reporting personnel, and staff turnover as primary obstacles to meeting weekly deadlines. For example, at health center O, there is a single health worker who is responsible for caring for patients, documenting clinical encounters, and reporting weekly surveillance data to the district biostatistician.

Furthermore, inadequate IDSR training among all health workers was noted as a challenge by personnel at these 3 facilities. While 2 staff members facilitate clinical activities at facility T, only 1 is trained to report data through mTrac. If the reporting person is absent from work, weekly data is not submitted. At health center D, staff turnover results in frequent loss of workers trained in IDSR and DHIS2 reporting, leaving behind personnel who are unfamiliar with reporting protocols. This is similar to facility H in Ngora, where staff members send photos of weekly 033b data to the reporting individual when he is absent from work as he is the only one with user rights to mTrac.

An additional obstacle described by personnel at 4 of the 7 health centers was the technological barrier posed by mTrac, as this program requires that reporting persons have access to cellular data or a wireless network. Participants at health centers O and T in Soroti City, and C and I in Soroti district, described unreliable access to cellular data, necessitating that they send pictures of the 033b report from their mobile phones to their respective district biostatisticians using the cellular application, WhatsApp. For those data which are not directly submitted via mTrac due to technical or other challenges, biostatisticians must manually enter the data into facility reports. The loss of cellular data access following withdrawal of aid by implementing partners (IPs) or changes in IP providers was also a common theme from key informant interviews. At three facilities (D and T in Soroti City and I in Soroti district), a lack of HMIS reporting tools was noted, including a lack of TB request forms and registries, laboratory request forms, and laboratory results report forms.

Minor themes from standardized interviews included delays in laboratory testing. At both health center D in Soroti City and H in Ngora district, study personnel spoke with participants who reported that delay in laboratory sample processing was the most significant barrier to timely 033b submission, stating that often samples are held at the nearby general hospital for two weeks before they are notified of results.

## DISCUSSION

Although improved health data reporting has been demonstrated across sub-Saharan Africa and within Uganda following DHIS2 implementation, we identified several obstacles to accurate, complete, and timely reporting of weekly surveillance data at regional, district, and health facility levels, with a number of districts and facilities within Teso consistently reporting below the national target of 80% for both metrics.^1,18^ Of the 11 reporting districts in the Teso sub-region, Soroti district, Soroti City, and Ngora district had the lowest rates of reporting completeness and timeliness over the 4 month study period. Studying the health facilities within these 3 districts more closely, we determined that the volume of reported outpatient visits was positively correlated with rates of 033b completeness in Soroti City. From our interviews with personnel at 7 different health centers across these three districts, we identified 5 predominant challenges to timely and complete reporting: (1) inadequate staffing due to financial constraints, (2) inadequate staff trained in data reporting and surveillance processes, (3) inconsistent access to cellular data or wireless internet, (4) inadequate HMIS reporting tools, and (5) laboratory delay in sample processing. Identification of these shared challenges allows for more targeted, standardized approaches to improving data reporting across not only single districts, but the whole country.

We hypothesized that higher volumes of outpatient visits at individual facilities would correlate with decreased rates of 033b reporting; however, there was no statistical significance between these variables for 2 of the districts (Soroti and Ngora). In Soroti City, there was a statistically significant positive correlation between OPD attendance and 033b reporting, meaning that higher patient volumes were associated with higher 033b reporting rates (completeness). We postulate that facilities with higher patient volumes are equipped with more human and material resources, facilitating more comprehensive and timely reporting.

Across health facilities, problems of understaffing as well as competing responsibilities and inadequate IDSR training among health workers appear to be significant challenges. Such challenges result in underreporting of priority disease events and, consequently, insufficient provision of resources needed to respond to such events when they occur.^6^ District surveillance officers often concomitantly manage numerous clinical and other duties, necessitating task-shifting.^6,19^ In many cases, patient care responsibilities are prioritized over data reporting, resulting in delayed, incomplete, or absent data submission at these centers.^6,20^

From our interviews, the problems of understaffing and competing responsibilities appear to be further exacerbated by the evident lack of personnel able or willing to assist with reporting– either due to staffing shortages or a lack of training– when designated reporters are too busy to report or absent.^11,21^ Because weekly statistics help inform responsible authorities of local priority disease trends supporting decisions at the points of generation, the lack of such data hinders decision-makers from allocating medical resources to districts in need, perpetuating problems of inequitable resource distribution and poor health outcomes in comparatively under-resourced localities.^11^ To respond to such issues and ensure shared responsibility when principal data reporting persons are indisposed, standardized skills training in IDSR and reporting protocols should be routine and mandatory among all staff members.^6,11,22^ Multiple studies conducted across Africa have demonstrated improved data reporting practices, specifically in the domains of timeliness and completeness, following IDSR training of health workers at both health facility and district levels.^8,20,22,23^ To ensure success, reporting tools must be readily available and feedback systems must be in place.^23^

Our findings further revealed problems of data accuracy and quality. At each level of the hierarchical reporting process, there is opportunity for error introduction due to both manual reporting practices in light of internet problems preventing the use of mTrac, frequent DHIS2 system failures affecting data entry, and lack of routine validation of priority disease statistics by supervisors prior to submission.^6,11,24^ In their review of barriers to the use of health data in low- and middle-income countries, authors Li et al. remark upon the disconnect between actors who produce data and those who use data for decision-making.^24^ From interviews with personnel, we came to understand that while many reporting persons are aware of DHIS2, few leverage it as a tool to analyze their own facility’s data or compare it to other health facilities within the district. By simply reporting data without validating it or understanding its direct impact on their health center or district, surveillance staff miss out on the true utility of the numbers they report.^11,24^ The limited interaction of data reporting persons with the data itself thus fosters mistrust of the data and a sense of unreliability on behalf of decision makers.^24^ Interestingly, at those facilities that were among the highest performing in their district, including health center C in Ngora, designated data officers frequently reviewed data with other staff members and displayed important trends within the facility. In order to ensure data fidelity and value, frequent validation of statistics with biostatisticians as well as encouragement of health facility staff to use data for local decision-making is of utmost importance.^6,18,20,23,24^

Nansikombi et al. previously studied rates of timely and complete health data reporting across Uganda from 2020-2021, postulating that inadequate training of health workers, limited supervision and mentorship, inadequate reporting tools, and network and internet challenges, among others, may contribute to low reporting rates.^6^ Our study further contextualizes and confirms such hypotheses through standardized in-person interviews with data reporting persons. The elucidation of barriers to accurate, complete, and timely reporting of weekly 033b data may facilitate more equitable, quality healthcare across not only Teso, but all of Uganda.

Our study had limitations. Firstly, we did not visit any privately operated health facilities, but only conducted interviews with health workers at government health centers. This is because of high rates of staff turnover, inconsistent government support, and reduced engagement with national reporting protocols in the private sector as such activities may compromise their business models.^6,18,22,25^ The lack of private facility participation in the nationwide reporting health database poses significant national and international problems when attempting to accurately assess the burden of epidemic prone diseases within Uganda.^25^ Beyond the limitations of accurate assessment, this lack of private facility input also poses a significant public health risk.^25^ Secondly, given human resource and time constraints, we visited only 7 of 241 reporting health facilities in Teso, reducing the generalizability of our qualitative findings from interviews with health personnel. Finally, because both OPD attendance and 033b data are reported by health personnel at individual facilities, it is possible that those facilities that underreport 033b data also underreport or fail to report monthly OPD attendance, which may further complicate our interpretation of the association between patient volumes and reporting rates.

## CONCLUSIONS

The implementation of DHIS2 throughout Uganda has had a dramatically positive impact upon public health and disease surveillance within the IDSR framework, but it has had challenges. It is important to identify and expound upon these challenges, as the value of DHIS2 lies in the comprehensiveness and validity of its data. Data timeliness and fidelity is key to actionable eHMIS data on subnational and national scales. Our mixed-methods analysis outlines key obstacles and friction points, some of which include inadequate surveillance staff at health facilities, insufficient IDSR trained health professionals, and lack of internet availability, that directly hinder health personnel from reporting timely, complete, and accurate data. While those issues related to inadequate financing and cost barriers may require more complex, multisectoral solutions, there are sustainable, well-studied approaches to addressing some of the challenges identified over the course of this study.^3,6,11^ With identification, targeted training and other interventions to relieve these obstacles and prevent public health disasters is possible.

## Data Availability

All data produced in the present work are contained in the manuscript

