## Supplemental Material for "Health Data Reporting Challenges in the Age of DHIS2: a Mixed-Methods Review in the Teso Sub-Region of Uganda"

**Supplementary Figure 1.** Standardized Questionnaire for Health Centers: Assessing Barriers to Timeliness and Completeness of 033b Data Collection & Reporting

1. What are the current challenges you face with regards to collecting and submitting routine surveillance data to DHIS2?
    a. Are there any challenges in particular that you believe this health center faces (ie staffing shortages etc)?
2. What are the reasons for delayed submission of data to DHIS2?
3. Are there any specific data elements that are more difficult to collect than others?

**Questions on review of the Outpatient Registers at poorly performing Health Facilities**

1. Are there data collection tools at the health facility including an Outpatient Department register?
2. Is complete data entered under the different headings?
3. Are dates of patient consult appended?
4. Are results of tests conducted appended for all conducted tests?
5. Is the suspected diagnosis appended for all patient entries?
6. Demographic variables
    a. Age is appended for all patient entries?
    b. Is gender appended for all the patient entries to the register?
    c. Is the patient address is appended for all patient entries to the register?

**Supplementary Table 1.** Rates of complete reporting (%) of 033b data by health centers in Soroti district, Uganda, January-April, 2024

| Soroti district | Monthly scores for completeness (%) | | | | Mean |
|---|---|---|---|---|---|
| Health Centers | January | February | March | April | |
| A (level II) | 100 | 100 | 75 | 100 | 94 |
| B (level II) | 100 | 100 | 50 | 100 | 88 |
| C (level IV) | 75 | 100 | 50 | 100 | 81 |
| D (level II) | 100 | 100 | 50 | 50 | 75 |
| E (level III) | 50 | 50 | 100 | 100 | 75 |
| F (level III) | 75 | 75 | 100 | 50 | 75 |
| G (level II) | 75 | 100 | 50 | 50 | 69 |
| H (level II) | 100 | 100 | 75 | 0 | 69 |
| I (level III) | 75 | 50 | 50 | 75 | 62 |
| J (level III) | 100 | 100 | 50 | 0 | 62 |
| K (level II) | 75 | 50 | 50 | 50 | 56 |
| L (level II) | 100 | 50 | 25 | 50 | 56 |
| M (level II) | 100 | 25 | 25 | 0 | 38 |
| N (level III) | 50 | 0 | 50 | 50 | 38 |
| O (level II) | 75 | 0 | 25 | 25 | 31 |
| P (level II) | 75 | 25 | 0 | 0 | 25 |
| **Median (IQR) aggregate score for Soroti district** | **--** | **--** | **--** | **--** | **65.5 (47-75)** |

**Supplementary Table 2.** Rates of timely reporting of 033b data by health centers in Soroti district, Uganda, January-April, 2024

| **Soroti district** | **Monthly scores for timeliness (%)** | | | | **Mean** |
|---|---|---|---|---|---|
| Health centers | January | February | March | April | |
| A (level II) | 100 | 100 | 75 | 75 | 88 |
| B (level II) | 75 | 100 | 50 | 50 | 69 |
| C (level IV) | 50 | 100 | 50 | 50 | 63 |
| D (level II) | 75 | 100 | 50 | 0 | 56 |
| E (level III) | 50 | 50 | 50 | 75 | 56 |
| F (level III) | 25 | 50 | 50 | 50 | 44 |
| G (level II) | 50 | 75 | 50 | 25 | 50 |
| H (level II) | 100 | 50 | 25 | 0 | 44 |
| I (level III) | 50 | 50 | 25 | 75 | 50 |
| J (level III) | 100 | 100 | 50 | 0 | 63 |
| K (level II) | 0 | 0 | 0 | 0 | 0 |
| L (level II) | 75 | 25 | 25 | 25 | 38 |
| M (level II) | 0 | 0 | 0 | 0 | 0 |
| N (level III) | 25 | 0 | 50 | 25 | 25 |
| O (level II) | 25 | 0 | 0 | 0 | 6 |
| P (level II) | 50 | 0 | 0 | 0 | 13 |
| **Median (IQR) aggregate score for Soroti district** | **---** | **---** | **---** | **---** | **47 (19-59.5)** |

**Supplementary Table 3.** Rates of complete reporting (%) of 033b data by health centers in Soroti City, Uganda, January-April, 2024

| Soroti City | Monthly scores for completeness (%) | | | | Mean |
|---|---|---|---|---|---|
| Health centers | January | February | March | April | |
| A (Regional Referral Hospital) | 100 | 100 | 100 | 100 | 100 |
| B (level III) | 75 | 100 | 100 | 100 | 94 |
| C (level III) | 75 | 100 | 100 | 100 | 94 |
| D (level III) | 100 | 100 | 100 | 75 | 94 |
| E (level III) | 75 | 100 | 100 | 100 | 94 |
| F (level III) | 75 | 100 | 100 | 75 | 88 |
| G (level III) | 75 | 75 | 100 | 100 | 88 |
| H* | 75 | 75 | 100 | 100 | 88 |
| I (level III) | 75 | 100 | 100 | 75 | 88 |
| J (level II) | 100 | 100 | 50 | 75 | 81 |
| K (level IV) | 75 | 50 | 100 | 100 | 81 |
| L (level III) | 100 | 100 | 50 | 75 | 81 |
| M (level II) | 75 | 50 | 100 | 100 | 81 |
| N (level II) | 50 | 50 | 100 | 100 | 75 |
| O (level II) | 100 | 100 | 50 | 50 | 75 |
| P (level III) | 75 | 100 | 50 | 75 | 75 |
| Q (level III) | 75 | 100 | 25 | 75 | 69 |
| R* | 100 | 75 | 25 | 50 | 63 |
| S* | 50 | 25 | 75 | 75 | 56 |
| T (level II) | 75 | 100 | 25 | 25 | 56 |
| U (level III) | 100 | 50 | 50 | 0 | 50 |
| V (level II) | 100 | 25 | 25 | 25 | 44 |
| W (level II) | 0 | 0 | 0 | 0 | 0 |
| X (level II) | 0 | 0 | 0 | 0 | 0 |
| Y* | 0 | 0 | 0 | 0 | 0 |
| **Median (IQR) aggregate score for Soroti City** | **--** | **--** | **--** | **--** | **81 (56-88)** |

*Unspecified facility level

**Supplementary Table 4.** Rates of timely reporting (%) of 033b data by health centers in Soroti City, Uganda, January-April, 2024

| **Soroti City** | **Monthly scores for timeliness (%)** | | | | **Mean** |
|---|---|---|---|---|---|
| Health centers | January | February | March | April | |
| A (Regional Referral Hospital) | 25 | 0 | 0 | 50 | 19 |
| B (level III) | 0 | 25 | 50 | 25 | 25 |
| C (level III) | 50 | 100 | 75 | 75 | 75 |
| D (level III) | 75 | 100 | 75 | 50 | 75 |
| E (level III) | 25 | 50 | 100 | 100 | 69 |
| F (level III) | 75 | 100 | 75 | 50 | 75 |
| G (level III) | 75 | 50 | 100 | 100 | 81 |
| H* | 50 | 50 | 25 | 75 | 50 |
| I (level III) | 75 | 100 | 75 | 50 | 75 |
| J (level II) | 100 | 100 | 50 | 50 | 75 |
| K (level IV) | 75 | 0 | 75 | 75 | 56 |
| L (level III) | 75 | 100 | 50 | 75 | 75 |
| M (level II) | 75 | 50 | 100 | 100 | 81 |
| N (level II) | 50 | 50 | 100 | 100 | 75 |
| O (level II) | 25 | 50 | 0 | 25 | 25 |
| P (level III) | 75 | 100 | 50 | 75 | 75 |
| Q (level III) | 50 | 100 | 25 | 75 | 63 |
| R* | 75 | 75 | 25 | 25 | 50 |
| S* | 50 | 25 | 50 | 50 | 44 |
| T (level II) | 75 | 75 | 0 | 25 | 44 |
| U (level III) | 100 | 50 | 50 | 0 | 50 |
| V (level II) | 100 | 25 | 25 | 25 | 44 |
| W (level II) | 0 | 0 | 0 | 0 | 0 |
| X (level II) | 0 | 0 | 0 | 0 | 0 |
| Y* | 0 | 0 | 0 | 0 | 0 |
| **Median (IQR) aggregate score for Soroti City** | **---** | **---** | **---** | **---** | **56 (34.5-75)** |

*Unspecified facility level

**Supplementary Table 5.** Rates of complete reporting of 033b data by health centers in Ngora district, Uganda, January-April, 2024

| **Ngora district** | **Monthly scores for completeness (%)** | | | | **Mean** |
|---|---|---|---|---|---|
| Health centers | January | February | March | April | |
| A (level III) | 100 | 100 | 50 | 75 | 81 |
| B (level III) | 100 | 100 | 75 | 50 | 81 |
| C (level III) | 100 | 100 | 50 | 75 | 81 |
| D* | 75 | 50 | 100 | 100 | 81 |
| E (level II) | 75 | 50 | 100 | 100 | 81 |
| F (level III) | 100 | 100 | 50 | 50 | 75 |
| G (level II) | 100 | 100 | 50 | 25 | 69 |
| H (level IV) | 25 | 75 | 75 | 75 | 63 |
| I (level III) | 100 | 75 | 50 | 0 | 56 |
| J (level II) | 100 | 100 | 25 | 0 | 56 |
| K (level III) | 100 | 75 | 25 | 0 | 50 |
| L (level II) | 25 | 0 | 0 | 0 | 6 |
| M (level II) | 25 | 0 | 0 | 0 | 6 |
| **Median (IQR) aggregate score for Ngora district** | **--** | **--** | **--** | **--** | **69 (53-81)** |

*Unspecified facility level

**Supplementary Table 6.** Rates of timely reporting of 033b data by health centers in Ngora district, Uganda, January-April 2024

| **Ngora district** | **Monthly scores for timeliness (%)** | | | | **Mean** |
|---|---|---|---|---|---|
| Health centers | January | February | March | April | |
| A (level III) | 50 | 75 | 50 | 75 | 63 |
| B (level III) | 75 | 100 | 25 | 50 | 63 |
| C (level III) | 100 | 100 | 50 | 75 | 81 |
| D* | 75 | 50 | 50 | 25 | 50 |
| E (level II) | 75 | 50 | 75 | 100 | 75 |
| F ((level III) | 100 | 100 | 50 | 50 | 75 |
| G (level II) | 75 | 75 | 25 | 25 | 50 |
| H (level IV) | 25 | 75 | 50 | 75 | 56 |
| I (level III) | 75 | 50 | 50 | 0 | 44 |
| J (level II) | 100 | 100 | 25 | 0 | 56 |
| K (level III) | 25 | 50 | 25 | 0 | 25 |
| L (level II) | 0 | 0 | 0 | 0 | 0 |
| M (level II) | 25 | 0 | 0 | 0 | 6 |
| **Median (IQR) aggregate score for Ngora district** | **---** | **---** | **---** | **---** | **56 (34.5-69)** |

*Unspecified facility level

**Supplementary Table 7.** Number of outpatient department visits per month by health facility in Soroti district, Uganda, January-April, 2024

| **Soroti district** | **Monthly number of OPD visits reported** | | | | **Total** |
|---|---|---|---|---|---|
| Health centers | January | February | March | April | |
| A (level II) | 244 | 354 | 197 | 245 | 1040 |
| B (level II) | 108 | 174 | None reported | 46 | 328 |
| C (level IV) | 1409 | 2131 | 771 | 1568 | 5879 |
| D (level II) | 554 | 543 | 546 | 45 | 1688 |
| E (level III) | 1490 | 1539 | 977 | 601 | 4607 |
| F (level III) | 1252 | 1378 | 1096 | 667 | 4393 |
| G (level II) | 400 | 472 | 217 | 123 | 1212 |
| H (level II) | 443 | 622 | 331 | None reported | 1396 |
| I (level III) | 916 | 1237 | 389 | 321 | 2863 |
| J (level III) | 1442 | 2135 | 820 | None reported | 4397 |
| K (level II) | 733 | 642 | 190 | 96 | 1661 |
| L (level II) | 623 | 350 | 124 | 283 | 1380 |
| M (level II) | 47 | None reported | 14 | None reported | 61 |
| N (level III) | 1297 | 874 | 718 | 880 | 3769 |
| O (level II) | 696 | 508 | 117 | 105 | 1426 |
| P (level II) | 224 | 152 | None reported | None reported | 376 |

**Supplementary Table 8.** Number of outpatient department visits per month by health facility in Ngora district, Uganda, January-April, 2024

| Ngora district | Monthly number of OPD visits reported | | | | Total |
|---|---|---|---|---|---|
| Health centers | January | February | March | April | |
| A (level III) | 1043 | 920 | 575 | 473 | 3011 |
| B (level III) | 1306 | 1908 | 658 | 206 | 4078 |
| C (level III) | 1732 | 1647 | 773 | 868 | 5020 |
| D* | 603 | 787 | 478 | 623 | 2491 |
| E (level II) | 273 | 190 | 84 | 119 | 666 |
| F (level III) | 1022 | 698 | 102 | 156 | 1978 |
| G (level II) | 320 | 310 | 80 | 80 | 790 |
| H (level IV) | 1805 | 2349 | 1745 | 2040 | 7939 |
| I (level III) | 671 | 387 | 200 | None reported | 1258 |
| J (level II) | 412 | 516 | 125 | None reported | 1053 |
| K (level III) | 942 | 803 | 336 | None reported | 2081 |
| L (level II) | 154 | None reported | None reported | None reported | 154 |
| M (level II) | 204 | None reported | None reported | None reported | 204 |

*Unspecified facility level

**Supplementary Table 9.** Number of outpatient department visits per month by health facility in Soroti City, Uganda, January-April, 2024

| **Soroti City** | **Monthly number of OPD visits reported** | | | | **Total** |
|---|---|---|---|---|---|
| Health centers | January | February | March | April | |
| A (Regional Referral Hospital) | None reported | 1138 | 4994 | 4739 | 10871 |
| B (level III) | 1013 | 1277 | 868 | 654 | 3812 |
| C (level III) | 969 | 1160 | 850 | 486 | 3465 |
| D (level III) | 510 | 862 | 387 | 555 | 2314 |
| E (level III) | 117 | 549 | 786 | 917 | 2369 |
| F (level III) | 786 | 1430 | 665 | 551 | 3432 |
| G (level III) | 1180 | 1165 | 891 | 1014 | 4250 |
| H* | 1075 | 2118 | 946 | 760 | 4899 |
| I (level III) | 1425 | 1249 | 1035 | 534 | 4243 |
| J (level II) | 159 | 422 | 259 | 277 | 1117 |
| K (level IV) | 1270 | 1989 | 1372 | 787 | 5418 |
| L (level III) | 111 | 96 | 64 | 63 | 334 |
| M (level II) | 125 | 141 | 81 | 98 | 445 |
| N (level II) | 147 | 104 | 145 | 173 | 569 |
| O (level II) | 395 | 383 | 182 | 95 | 1055 |
| P (level III) | 249 | 358 | 167 | 219 | 993 |
| Q (level III) | 23 | 54 | 8 | 41 | 126 |
| R * | 745 | 881 | 725 | 763 | 3114 |
| S* | 6 | 12 | 14 | 16 | 48 |
| T (level II) | 277 | 415 | 103 | 29 | 824 |
| U (level III) | 225 | 84 | 68 | None reported | 377 |
| V (level II) | 452 | 81 | 109 | 77 | 719 |
| W (level II) | None reported | None reported | None reported | None reported | –- |
| X (level II) | None reported | None reported | None reported | None reported | –- |
| Y* | None reported | None reported | None reported | None reported | –- |

*Unspecified facility level
